# A betacoronavirus multiplex microsphere immunoassay detects early SARS-CoV-2 seroconversion and controls for pre-existing seasonal human coronavirus antibody cross-reactivity

**DOI:** 10.1101/2020.10.14.20207050

**Authors:** Eric D. Laing, Spencer L. Sterling, Stephanie A. Richard, Shreshta Phogat, Emily C. Samuels, Nusrat J. Epsi, Lianying Yan, Nicole Moreno, Christian Coles, Jennifer Mehalko, Matthew Drew, Caroline English, Kevin K. Chung, G. Travis Clifton, Vincent J. Munster, Emmie de Wit, David Tribble, Brian K. Agan, Dominic Esposito, Charlotte Lanteri, Edward Mitre, Timothy H. Burgess, Christopher C. Broder

## Abstract

With growing concern of persistent or multiple waves of SARS-CoV-2 in the United States, sensitive and specific SARS-CoV-2 antibody assays remain critical for community and hospital-based SARS-CoV-2 surveillance. Here, we describe the development and application of a multiplex microsphere-based immunoassay (MMIA) for COVD-19 antibody studies, utilizing serum samples from non-human primate SARS-CoV-2 infection models, an archived human sera bank and subjects enrolled at five U.S. military hospitals. The MMIA incorporates prefusion stabilized spike glycoprotein trimers of SARS-CoV-2, SARS-CoV-1, MERS-CoV, and the seasonal human coronaviruses HCoV-HKU1 and HCoV-OC43, into a multiplexing system that enables simultaneous measurement of off-target pre-existing cross-reactive antibodies. We report the sensitivity and specificity performances for this assay strategy at 98% sensitivity and 100% specificity for subject samples collected as early as 10 days after the onset of symptoms. In archival sera collected prior to 2019 and serum samples from subjects PCR negative for SARS-CoV-2, we detected seroprevalence of 72% and 98% for HCoV-HKU1 and HCoV-0C43, respectively. Requiring only 1.25 µL of sera, this approach permitted the simultaneous identification of SARS-CoV-2 seroconversion and polyclonal SARS-CoV-2 IgG antibody responses to SARS-CoV-1 and MERS-CoV, further demonstrating the presence of conserved epitopes in the spike glycoprotein of zoonotic betacoronaviruses. Application of this serology assay in observational studies with serum samples collected from subjects before and after SARS-CoV-2 infection will permit an investigation of the influences of HCoV-induced antibodies on COVID-19 clinical outcomes.

## INTRODUCTION

Severe acute respiratory syndrome coronavirus-2 (SARS-CoV-2) is a novel zoonotic positive-sense, single-stranded, RNA virus responsible for the third viral pandemic of the 21^st^ century, and the third zoonotic coronavirus outbreak in the past 20 years (1, 2). At this time, SARS-CoV-2 has globally caused 34 million COVID-19 cases and over 1 million COVID-19 related deaths. A major concern of the ongoing SARS-CoV-2 pandemic has been the frequent reports of waning virus-specific antibody levels, with several studies reporting decay to undetectable levels within just a few months after infection (3-5). While this is a measurable feature of antibody response, it is also possible that current assays lack the sensitivity required to detect lower levels of SARS-CoV-2 specific antibodies. To date, a variety of antibody tests have been developed with 38 tests granted Emergency Use Authorization (EUA) by the U.S. Food and Drug Administration. The majority of these tests assess for antibodies against the coronavirus spike (S) envelope glycoprotein, the primary target of virus-neutralizing antibodies (6), in either its native-like oligomer conformation, or against one of its protein subunits or domains. In general, most S glycoprotein antigen-based assays report the ability to detect antibodies in 65-70% of infected individuals 8 – 14 days after symptom onset, with positivity rates over 90% not occurring until 2 – 3 weeks after symptom onset (7).

In this study, we describe the development, characterization, and utility of a betacoronavirus (β-CoV) multiplex microsphere-based immunoassay (MMIA) for COVID-19 serology studies. To optimize sensitivity and specificity for measuring SARS-CoV-2 spike reactive antibodies, the MMIA included prefusion stabilized S glycoprotein ectodomain trimers of SARS-CoV-2, SARS-CoV-1, MERS-CoV, and the seasonal human coronaviruses (HCoV), HCoV-HKU1 and HCoV-OC43. The MMIA enabled the simultaneous measurement of relative antibody quantities against each of these medically-relevant betacoronaviruses. We hypothesized that this approach would potentially result in a highly sensitive and specific assay for detecting SARS-CoV-2 specific antibodies through two mechanisms. First, the Luminex xMAP-based platform has a large dynamic range and has been shown to be more sensitive than ELISA for the detection of antibodies to other viral infections (8-10). Second, given the high seroprevalence of the common human betacoronaviruses (11-13), cross-reactive antibodies present in subject samples (14, 15) could be concurrently measured and accounted for in a multiplex approach. By testing for S glycoprotein reactive antibodies to SARS-CoV-2 in the presence of HKU1 and OC43 S glycoproteins, the MMIA assay controls for off-target pre-existing cross-reactive betacoronavirus antibodies, thus enhancing specific SARS-CoV-2 antibody detection. Additionally, the simultaneous incubation of serum with S glycoproteins from all the relevant betacoronaviruses may enable a lower threshold for SARS-CoV-2 antibody positivity.

Utilizing serum samples from an experimentally challenged non-human primate (NHP) model, together with human sera from subjects confirmed to have SARS-CoV-2 infection and from subjects confirmed to have other coronavirus infections collected prior to 2018, we report the sensitivity and specificity performances for this assay strategy. Serum samples from rhesus macaques experimentally infected with SARS-CoV-2 demonstrated that SARS-CoV-2 S glycoprotein IgG seroconversion was detectable by 10 days post infection (dpi), consistent with other reports demonstrating anti-S glycoprotein IgG seroconversion between 3 and 14 dpi (16-19). As a result, we evaluated serum samples from SARS-CoV-2 positive subjects collected 10 days after symptom onset and report 98% sensitivity for SARS-CoV-2 S glycoprotein IgG antibody detection in humans at that time point. We also examined differences in SARS-CoV-2 antibody reactivity between widely used antigens: SARS-CoV-2 prefusion stabilized S glycoprotein ectodomain trimer and the receptor-binding domain (RBD). High seroprevalence of seasonal HCoV OC43 and HKU1, ranging from 97 – 98% and 55 – 89%, respectively, was observed across both archival sera and SARS-CoV-2 negative subject serum samples. Through this MMIA strategy we aim to investigate the interplay of pre-existing seasonal HCoV antibodies on SARS-CoV-2 IgG duration, COVID-19 symptom presentation, and disease severity. Preliminary data we have obtained using this multiplex serology strategy demonstrates that SARS-CoV-2 antibody can be detected be detected early after the onset of symptoms and that SARS-CoV-2 infection can stimulate an IgG antibody response that is cross-reactive with SARS-CoV-1 and MERS-CoV S glycoproteins.

## RESULTS

### Comparison of MMIA and ELISA for SARS-CoV-2 IgG antibody detection

We first established our ability to detect SARS-CoV-2 IgG and monitor SARS-CoV-2 seroconversion with sera collected from SARS-CoV-2 infected NHP. Purified IgG from SARS-CoV-2 infected NHP collected 21 dpi were pooled and spiked into NHP negative sera, and the MMIA was quantitatively characterized for IgG polyclonal reactivity revealing SARS-CoV-2 spike antibody MFI curve linearity between 0.625 – 5.0 µg/ml or 3690 – 20,354 MFI (Figure 1). Positive MMIA saturation occurs within 20,000 – 30,000 MFI. To investigate the effects of the increased dynamic range facilitated by Luminex xMAP-based multiplexing systems on MMIA sensitivity, we compared end-point titers by both ELISA and MMIA. In an ELISA, SARS-CoV-2 positive NHP sera end-point titers ranged from 1,000 to 2,000 (Figure 2A), consistent with reported ELISA titers for these animals (16). In the MMIA, the ability to detect serially diluted IgG antibodies was 4-to 8-fold greater than ELISA with end-point titers ranging from 4,000 (n= 2) to 16,000 (n= 1) (Figure 2B).

**Figure 1.**
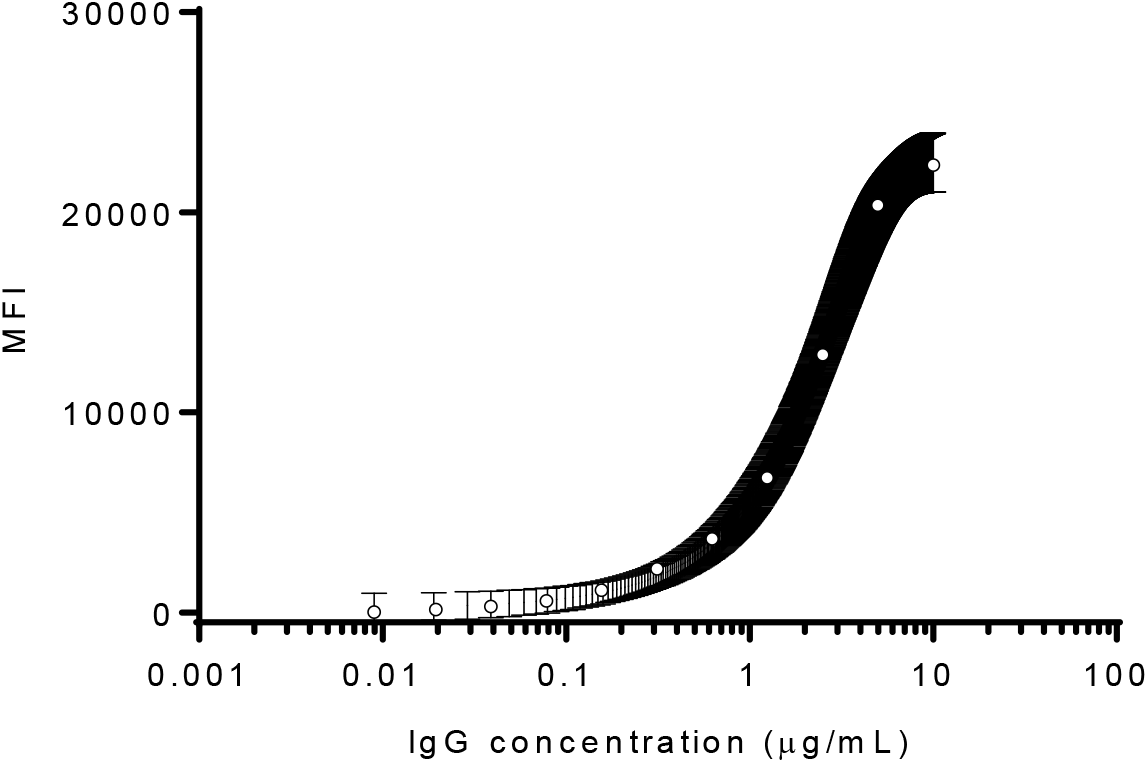
SARS-CoV-2 spike protein reactivity as a function of IgG concentration. A sigmoidal curve was used to fit the MEAN±SEM of two independent experiments performed in technical triplicates. MFI, median fluorescence intensities.

**Figure 2.**
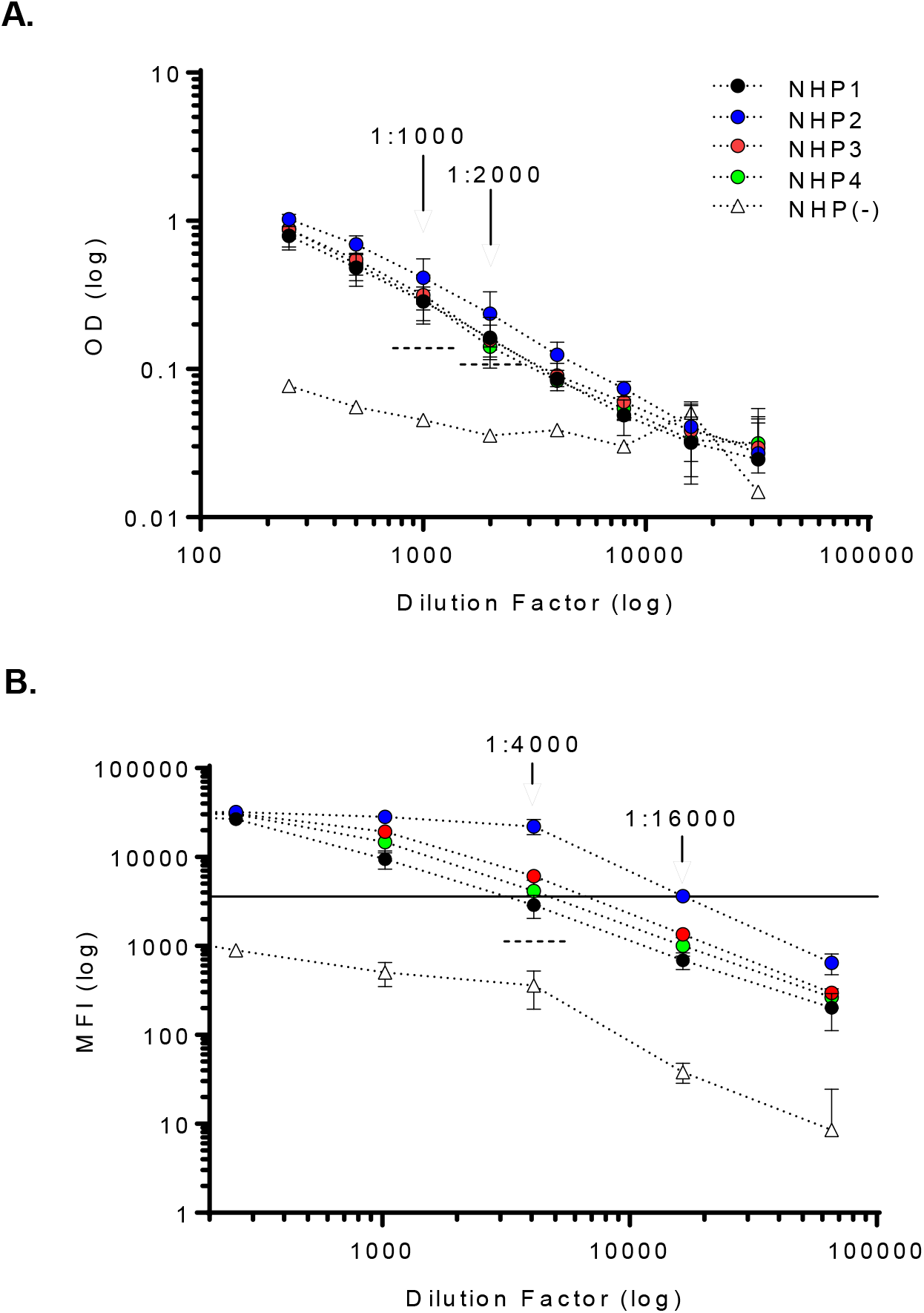
MMIA displays enhanced sensitivity for SARS-CoV-2 IgG detection. Serum samples from SARS-CoV-2 infected NHP collected 21 dpi were tested by SARS-CoV-2 spike protein **(A)** ELISA and **(B)** MMIA. **(A)** Dashed lines indicate 3-fold MFI above the NHP(-) serum sample(s) diluted 1:1000 and 1:2000. **(B)** A solid line indicates the lower limit of MFI linearity and a dashed line indicates 3-fold MFI above the NHP(-) serum sample diluted 1:4000. Positive samples are those above both the MFI level for curve linearity and 3-fold change in NHP(-) serum. MFI values represent MEAN±SD of two independent experiments performed in technical triplicates.

### Seroconversion in a non-human primate model

Next, we monitored SARS-CoV-2 seroconversion with longitudinal NHP serum samples. SARS-CoV-2 spike protein reactive IgG antibody seroconversion was observed in all four NHP 10 dpi (Figure 3A). We also investigated SARS-CoV-2 IgM antibody seroconversion and detected IgM level above baseline in two NHP by 7 dpi; all four NHP had detectable IgM by 10 dpi (Figure 3B). Notably, IgG antibody from SARS-CoV-2 challenged NHP did not significantly react with spike proteins from betacoronaviruses, SARS-CoV-1, MERS-CoV or HCoVs, included in the MMIA (Figure 3A), whereas, a varying degree of IgM cross-reactivity was observed (Figure 3B). Additionally, high baseline IgM reactivity to SARS-CoV-2 RBD at 0 dpi inhibited our ability to ascertain the dpi where seroconversion could be observed with this SARS-CoV-2 antigen (Figure 3B).

**Figure 3.**
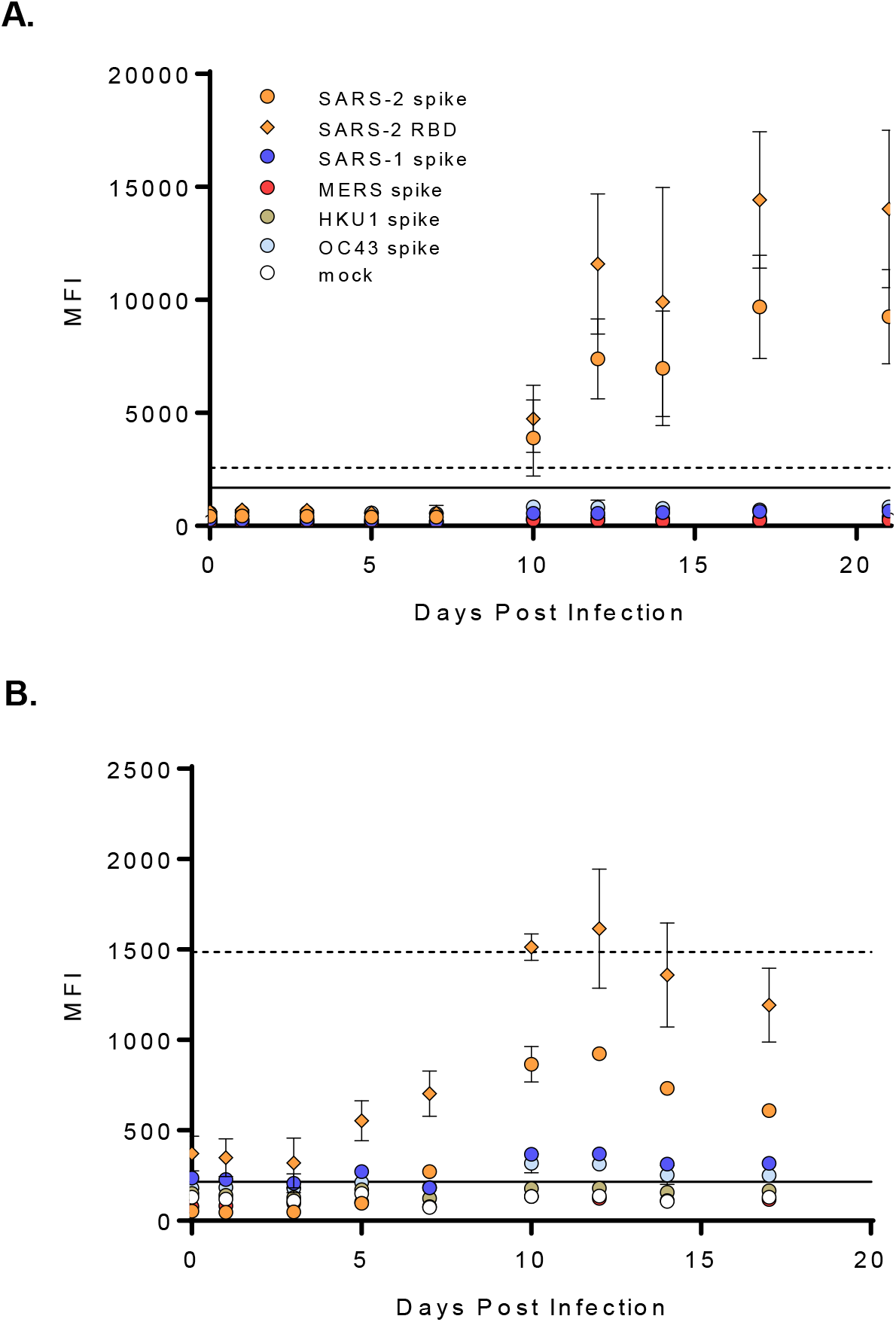
SARS-CoV-2 seroconversion in a non-human primate model. Sera from SARS-CoV-2 infected NHP were screened for SARS-CoV-2 spike protein reactive **(A)** IgG and **(B)** IgM. Graphs represent the MEAN±SD of four SARS-CoV-2 challenged NHP screened in two independent experiments performed in technical duplicates. A solid line indicates a 4-fold rise in SARS-CoV-2 spike protein MFI from baseline (0 dpi) and was used as a threshold cutoff for SARS-CoV-2 IgG and IgM seroconversion; a dashed line indicates a 4-fold rise in SARS-CoV-2 RBD protein MFI from baseline.

### Archival sera from subjects with PCR-confirmed seasonal coronaviruses exhibit cross-reactivity with SARS-CoV-2 spike protein

Despite low sequence similarity and identity between SARS-CoV-2 spike protein and seasonal HCoV spike proteins, antibody cross-reactivity with SARS-CoV-2 proteins has been observed (14, 15). To determine whether prior infection with seasonal HCoV induces antibodies that cross-react with SARS-CoV-2, we assayed archival (pre-2019) serum from human subjects with PCR-confirmed seasonal HCoVs. When setting a cut-off for positivity at three times the mean MFI obtained for a mock antigen preparation-coupled microsphere, we observed that 8.89% (4/45) of archived serum samples from HCoV PCR-positive subjects cross-reacted with SARS-CoV-2 spike protein (Figure 4A-D). Cross-reactivity between HCoV-induced antibodies with SARS-CoV-2 spike protein was observed in subjects that were PCR-positive for OC43 (1/16), HKU1 (1/6) and 229E (2/10) infection.

**Figure 4.**
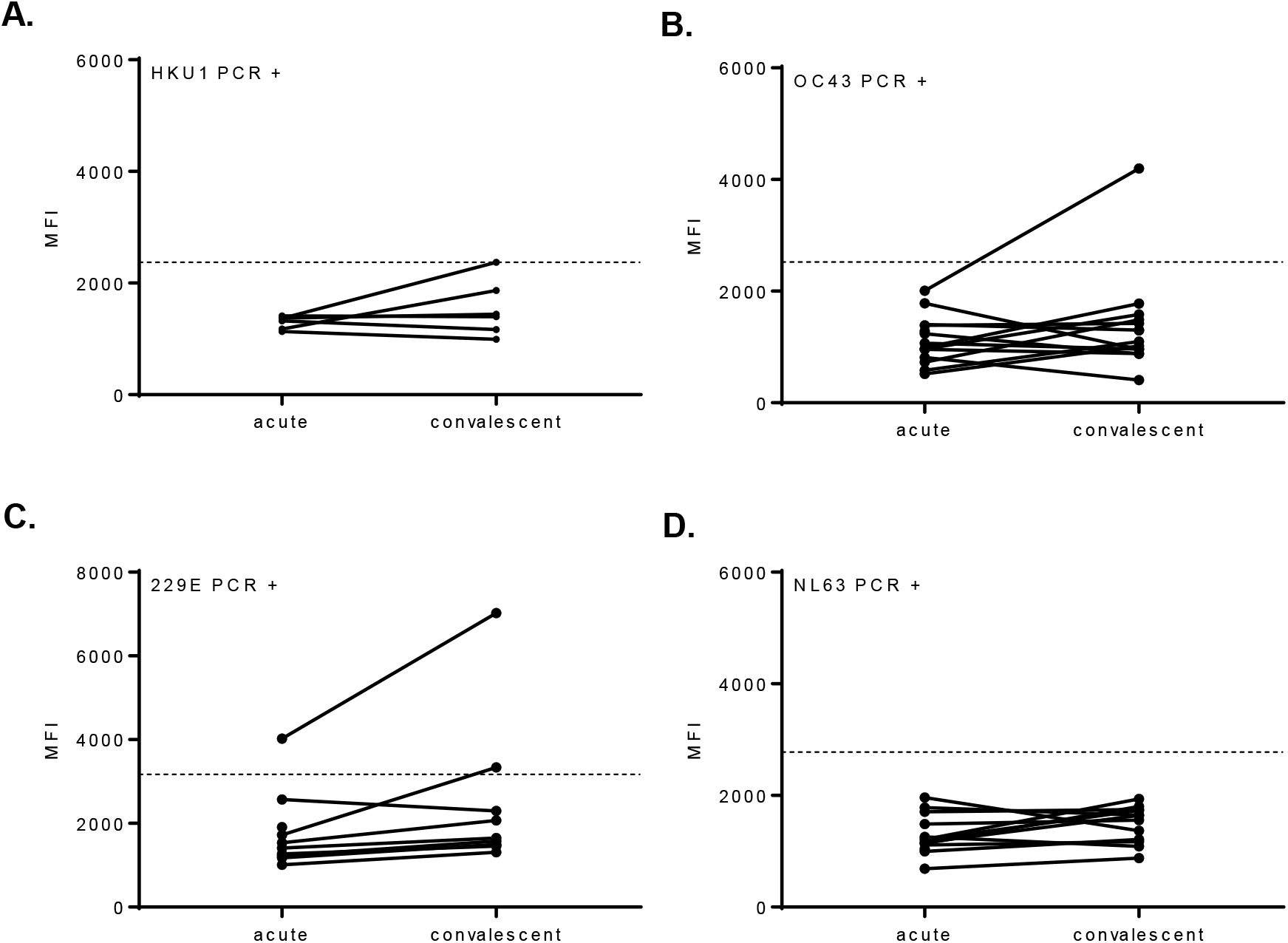
Archival sera from subjects with seasonal HCoVs can display cross-reactivity with SARS-CoV-2 spike protein. Acute and convalescent serum samples from HCoV PCR-positive subjects were tested in a β-CoV MMIA. Subjects are grouped together based on HCoV PCR confirmation, **(A)** OC43 (n= 16), **(B)** HKU1 (n= 6), **(C)** NL63 (n= 13) and **(D)** 229E (n= 10), A dashed line indicates the 3-fold change in the mean MFI of a mock antigen-coupled microsphere. MFI values represent the MEAN of two independent experiments performed in technical duplicates. MFI, median fluorescence intensity.

### Assay threshold cutoffs for SARS-CoV-2 spike protein reactive antibodies

To control for pre-existing HKU1 and OC43 spike protein reactive antibody cross-reactivity with SARS-CoV-2 spike protein, rather than using a cutoff of three times the mean MFI obtained for mock antigen, antibody threshold cutoffs were established with HCoV PCR-positive convalescent sera (Figure 5A-B). A conventional 99.7% probability, mean and three standard deviations higher, threshold cutoff was employed to distinguish positive and negative IgG antibodies. The mean IgG reactivity to SARS-CoV-2 spike protein was 1569 MFI with a 99.7% probability threshold cutoff of 4911 MFI. The SARS-CoV-2 RBD protein had a notably higher background IgG antibody reactivity and threshold cutoff, 2846 MFI and 7951 MFI, respectively. Given the inherently less-specific nature of IgM, a 99.9% probability threshold cutoff was preferred. The 99.9% probability threshold cutoffs for SARS-CoV-2 spike protein and RBD protein reactive IgM were 846 MFI and 15352 MFI. The remaining archival sera (n= 84), representing HCoV PCR-positive acute sera, rhinovirus PCR-positive acute/convalescent sera, and acute/convalescent sera from ‘no pathogen detected’ subjects, did not react with SARS-CoV-2 spike protein or RBD protein above the established threshold cutoffs for either antigen (Figure 5C-D). Although only 17%/7% of ARIC human subjects were OC43/HKU1 PCR positive, we observed 97.6% OC43 and 89.2% HKU1 IgG, and none were IgM positive (Figure 5C-D). Interestingly, the three OC43 spike IgM positive serum samples were collected from subjects who had no pathogen detected by PCR.

**Figure 5.**
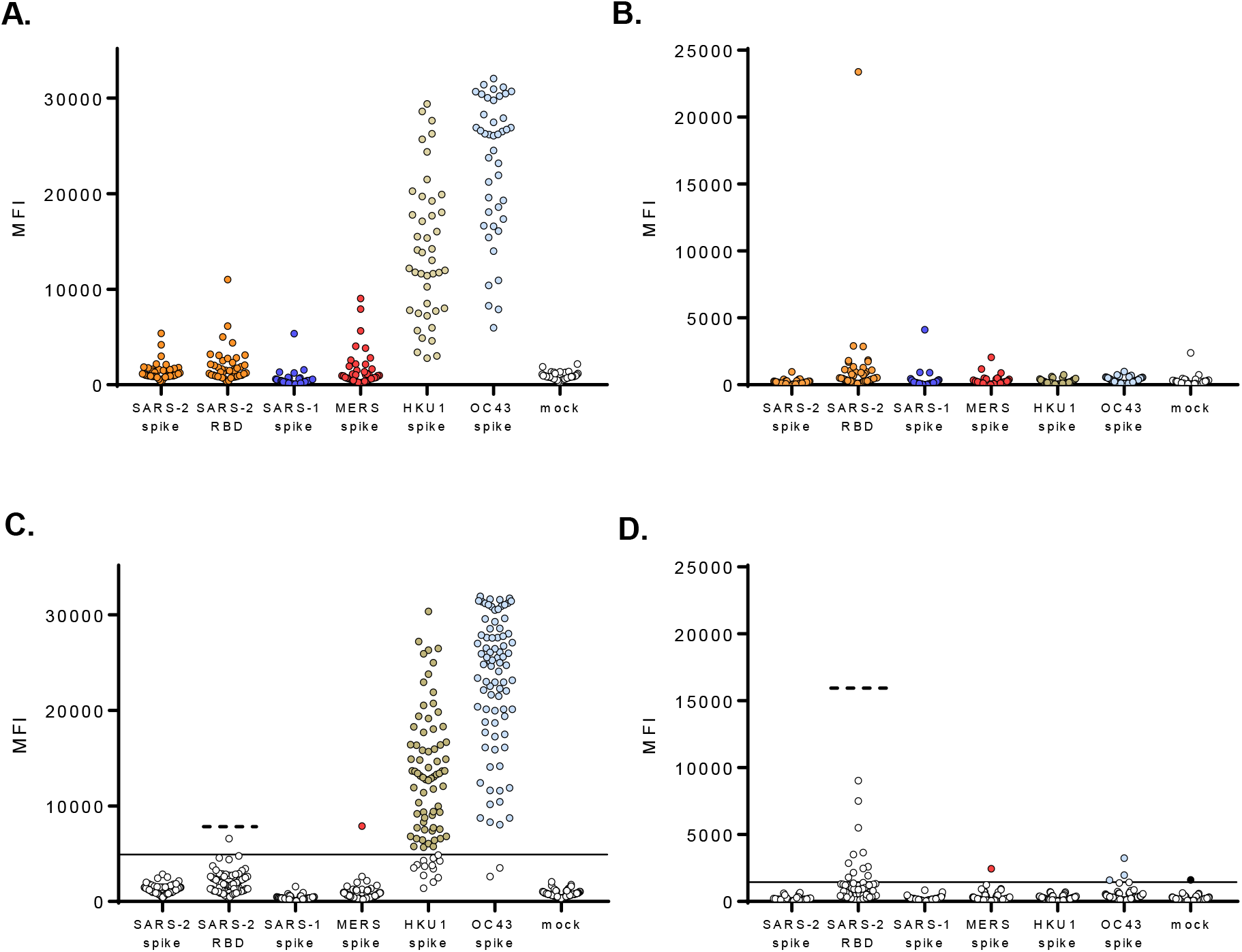
Archival sera generated threshold cutoffs for IgG and IgM antibodies confer specificity for SARS-CoV-2. Convalescent serum samples (n= 43) from HCoV PCR-positive subjects were tested in the β-CoV MMIA with **(A)** IgG antibody and **(B)** IgM antibody. **(C-D)** Archival sera (n= 84), acute serum samples from HCoV PCR-positive subjects, acute/convalescent serum samples from rhinovirus PCR-positive subjects and acute/convalescent serum samples from ‘no pathogen detected’ subjects were tested for **(C)** IgG and **(D)** IgM antibody reactivity tested against the established threshold cutoffs. A solid line indicates the threshold cutoff for positivity with SARS-CoV-2 spike protein and a dashed line indicates the threshold cutoff for SARS-CoV-2 RBD. Colored dots in **(C-D)** indicate samples with MFI above the SARS-CoV-2 spike protein threshold cutoff for positive antibody. MFI is the average of sera diluted 1:400, adjusted with PBS controls and tested across technical duplicate plates. IgG data is a representation of three independent screenings.

### Multiplex microsphere-based immunoassay performance

Sera from persons with SARS-CoV-2 infection were screened for IgG and IgM antibody reactivity with betacoronavirus spike proteins in our MMIA. As SARS-CoV-2 seroconversion was detected by 10 dpi in NHPs, MMIA SARS-CoV-2 spike protein IgG sensitivity for human serology was evaluated in confirmed subjects ≥ 10 days post-symptom onset (dpso). Additionally, a combination of archival human serum samples, and PCR negative MTF hospitalized subjects and outpatients with serum samples collected < 30 dpso were included in a negative agreement analysis. Subjects that were PCR confirmed as SARS-CoV-2 negative displayed no IgG with SARS-CoV-2 spike protein, whereas one subject had IgM reactivity to SARS-CoV-2 spike protein above the 99.9% cutoff MFI value. A receiver operating characteristic (ROC) curve analysis was then performed to further establish a more conservative threshold value (1446 MFI cutoff point) for SARS-CoV-2 positive IgM (Figure 6A-B). Further, HKU1 and OC43 IgG antibodies were observed in SARS-CoV-2 PCR negative subjects (Figure 6A). SARS-CoV-2 reactive IgG antibodies were only detected above the 99.7% threshold cutoff in the PCR positive subjects enrolled at military hospitals or the Javits Center field hospital; cross-reactive antibodies to SARS-CoV-1 and MERS-CoV were observed in SARS-CoV-2 PCR positive and IgG positive subjects (Figure 6B).

**Figure 6.**
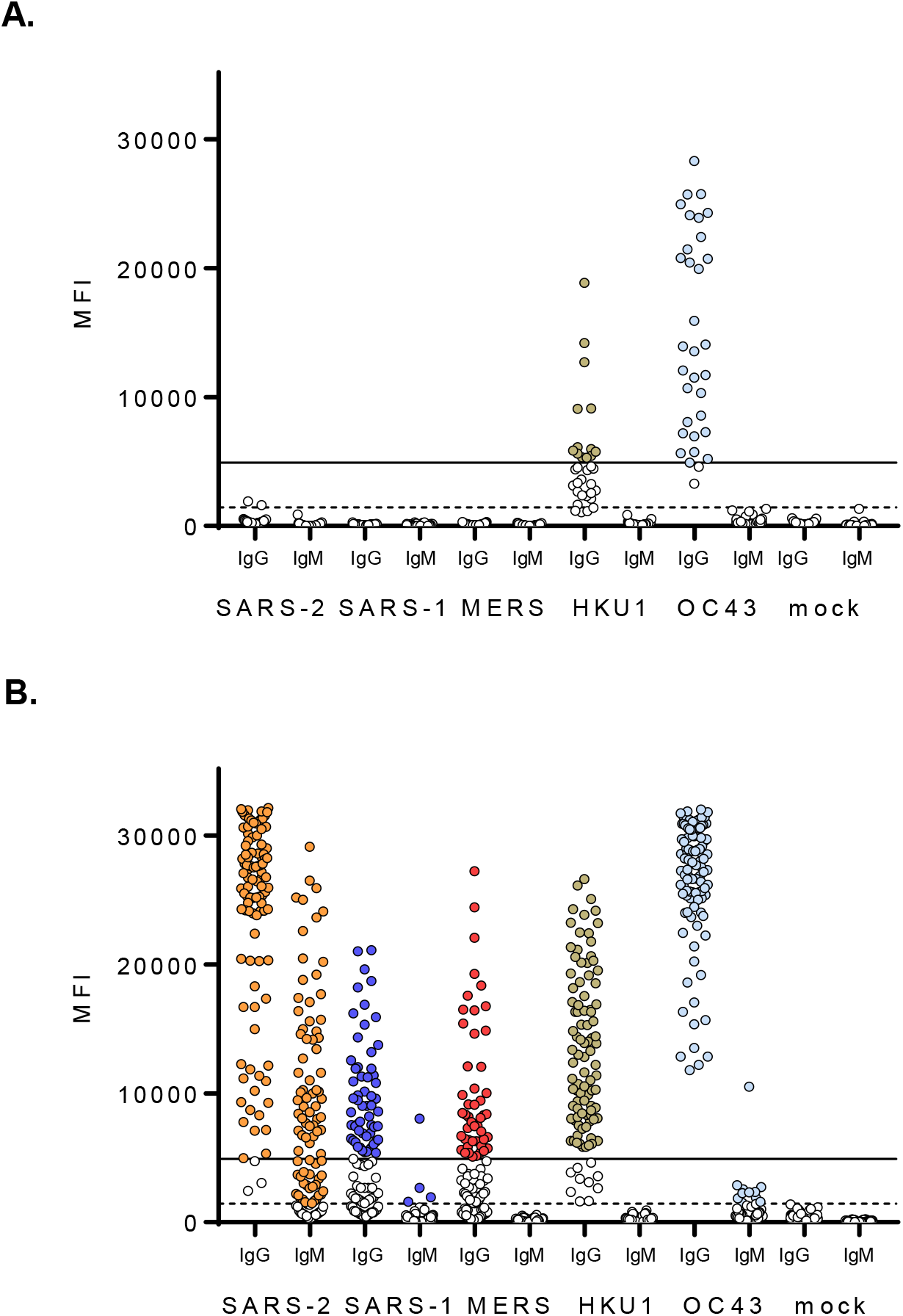
A multiplex antibody test can detect SARS-CoV-2 specific and cross-reactive antibodies. Serum samples from **(A)** SARS-CoV-2 PCR negative subjects **(B)** SARS-CoV-2 PCR-positive subjects collected ≥ 10 dpso were tested by β-CoV MMIA. Serum were diluted 1:400 and tested in duplicate plates. MFI, median fluorescence intensities, are the average of PBS-subtracted technical duplicates. A solid line indicates the IgG threshold cutoff and a dashed line indicates the IgM threshold cutoff. Colored dots indicate positive serum samples. SARS-2, SARS-COV-2; SARS-1, SARS-CoV-1; MERS, MERS-CoV; HKU1, HCoV-HKU1; OC43, HCoV-OC43.

Performance assessments of this β-CoV MMIA for SARS-CoV-2 spike protein IgG and IgM detection are included in Table 1. SARS-CoV-2 spike protein reactive IgG antibody detection was calculated with 95% confidence intervals (CI) as follows, sensitivity = 98.06% (94.45% – 99.60% CI), specificity= 100% (96.82% – 100.00% CI). Assuming a US disease prevalence of 1.0%, the positive predictive value (PPV) = 100.00%, and negative predictive value (NPV) = 99.98% (99.94% - 99.99% CI). SARS-CoV-2 spike protein reactive IgM detection sensitivity was lower than IgG, with performance analysis conducted with serum samples collected ≥ 7 days post-symptom onset, sensitivity= 78.10% (68.97% to 85.58%), specificity= 100% (96.95% - 100.00% CI) and PPV= 100.00% (Table 1).

**Table 1.** MMIA SARS-CoV-2 spike protein performance

|  | SARS-CoV-2 PCR Status/Archival Sera |  |  |  |
| --- | --- | --- | --- | --- |
| SARS-CoV-2<br>MMIA IgG<br>Antibody Test<sup>2</sup> |  | Positive<sup>1</sup> | Negative | Total |
|  | Positive | 152 | 0 | 152 |
|  | Negative | 3 | 117 | 120 |
|  | Total | 155 | 117 | 272 |
|  | Sensitivity | 98.1% |  |  |
|  | Specificity | 100% |  |  |
| SARS-CoV-2<br>MMIA IgM<br>Antibody Test<sup>3</sup> | Positive | 82 | 0 | 82 |
|  | Negative | 23 | 117 | 140 |
|  | Total | 105 | 117 | 222 |
|  | Sensitivity | 78.1% |  |  |
|  | Specificity | 100% |  |  |

<sup>1</sup>84 archival serum samples and 33 serum samples from PCR negative study enrollees are included as SARS-CoV-2 negative.
<sup>2</sup>IgG antibody test included serum samples from n= 155 PCR positive subjects including 82 EPICC outpatients, 38 EPICC hospitalized subjects and 35 COVID-19 NYC hospitalized subjects
<sup>3</sup>IgM antibody test included serum samples from n= 105 PCR positive subjects including 41 EPICC outpatients, 29 EPICC hospitalized subjects and 35 COVID-19 NYC hospitalized subjects

When comparing spike protein and RBD, notably, utility of SARS-CoV-2 RBD for IgG detection had a reduced sensitivity (87.10%, 80.78% - 91.94% CI) (Table 2). To assess MMIA precision, three positive and one negative samples were tested over five independent experiments, with at least two distinct in-house antigen-coupled bead lots and serum sample freeze-thaws. Coefficient of variations (CV) were calculated for all three positive samples and remained <20% (Figure 7). Although the negative sample had a > 20%, the MFI never went above the threshold cutoff for positive IgG across five independent tests.

**Table 2.** MMIA SARS-CoV-2 RBD performance

|  | SARS-CoV-2 PCR Status/Archival Sera |  |  |  |
| --- | --- | --- | --- | --- |
|  |  | Positive | Negative | Total |
| SARS-CoV-2<br>MMIA IgG<br>Antibody Test | Positive | 135 | 0 | 135 |
|  | Negative | 20 | 117 | 137 |
|  | Total | 155 | 117 | 266 |
|  | Sensitivity | 87.1% |  |  |
|  | Specificity | 100% |  |  |

**Figure 7.**
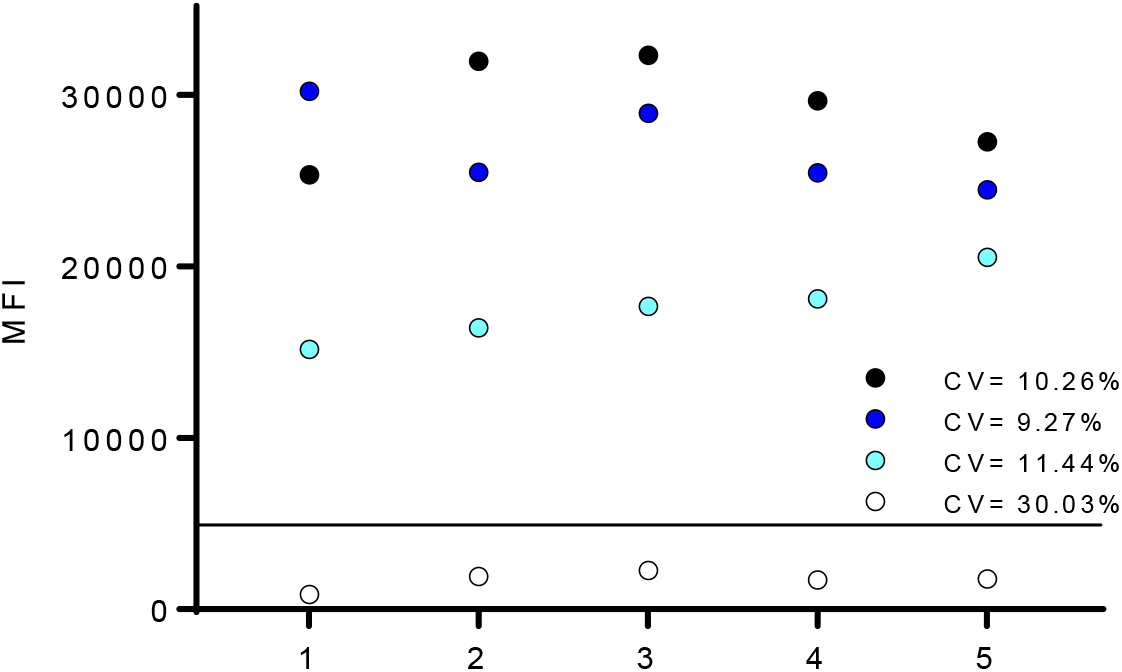
Positive and negative results are reproducible over independent MMIA tests. Selected positive(s) and negative serum samples were tested across independent experiments. CV, coefficient of variation, percentages are indicated on the graphs for each sample. A solid line indicates the threshold cutoff for positive IgG.

## DISCUSSION

In this study, we have demonstrated that use of a multiplex microsphere-based immunoassay (MMIA) built using Luminex xMAP-based technology in which individual microspheres are bound to pre-fusion stabilized S glycoprotein trimers of SARS-CoV-2, SARS-CoV-1, MERS-CoV, and each of the two seasonal betacoronaviruses enables highly sensitive and specific detection of SARS-CoV-2 IgG antibodies. In contrast to commercial ELISA and lateral flow assays for SARS-CoV-2 IgG, which typically have a sensitivity in the range of 65-70% up to 14 days after symptom onset (7), the MMIA has a sensitivity of 98% at just 10 days after symptom onset in PCR-confirmed cases of SARS-CoV-2 infection. Use of this highly sensitive assay will allow for improved assessments of the kinetics of humoral responses to SARS-CoV-2 infection.

We hypothesize that the high sensitivity and specificity of the MMIA assay is due both to the physics of the Luminex xMAP-based platform, which enables a high dynamic range of measurement, as well as to the use of a multiplexing system. Multiplex microsphere-based immunoassays have been shown to be more sensitive than standard ELISA for SARS-CoV-2 antibody detection (20) and several other virus infections, including Lassa virus, Ebola virus, and simian immunodeficiency virus (8-10). Additionally, and perhaps more importantly, simultaneously incubating serum against spike proteins of seasonal HCoVs may enable the establishment of a lower threshold of positivity for detection of SARS-CoV-2 antibodies. Given the presence of cross-reactive antibodies, assays that only test for antibodies against SARS-CoV-2 may have to utilize a high signal threshold as a cut-off for positivity to reduce false positive rates of detection. By incubating serum against multiple coronavirus spike proteins, the MMIA platform may allow preferential binding of cross-reactive antibodies to the antigens of the coronavirus against which they were initially induced, enabling a lower, and potentially more sensitive, cut-off for the detection of SARS-CoV-2 specific antibodies.

Now nine months into the COVID-19 pandemic, this β-CoV MMIA adds to an established body of antibody tests and serology results. Early in the pandemic, SARS-CoV-2 IgG seroconversion was surprisingly detected early after exposure and sometimes in parallel with IgM seroconversion (21-23). The temporal window to capture SARS-CoV-2 IgM is shorter than IgG, and with IgG seroconversion occurring 10 dpi in NHP SARS-CoV-2 disease models, and detectable as early as 7 dpso in subjects, there appears little benefit for continued SARS-CoV-2 IgM detection (Table 3). As we placed no upper limit on the dpso of the first serum collection included in performance analysis, IgM sensitivity is lower than IgG driven by outpatient enrollments in the EPICC, IDCRP-085 study on average 28 dpso that were IgG positive, but IgM negative. Although less sensitive and less specific than IgG detection, the benefit of IgM detection may lay in its ability to place a temporal window on SARS-CoV-2 exposure in asymptomatic IgG positive individuals, particularly useful for cross-sectional studies of seroprevalence.

**Table 3.** IgG and IgM seropositivity within 28 days post-symptom onset (dpso)

| dpso | IgG+ | IgG+/IgM+ |
| --- | --- | --- |
| 7 – 14 | 80.0% (12/15) | 73.3% (11/15) |
| 15 – 28 | 100% (31/31) | 93.5% (29/31) |

Conservation of epitopes present in the prefusion stabilized native-like trimeric S glycoprotein oligomers may be the major factor in the observed cross reactions between the CoV S glycoproteins. Since the RBD protein is only a domain within the S1 subunit of the S glycoprotein and lacks potentially conserved protein residues with seasonal HCoV S glycoproteins, utility in antigen-based immunoassays confers specificity for SARS-CoV-2 and is thus employed in several antibody tests (24-27). We noted that in the present β-CoV MMIA, the commercially sourced RBD protein, which shares an equivalent number of protein residues with the expressed RBD protein used in a microsphere-based immunoassay developed by the Ragon Institute of MGH, MIT and Harvard (20), had a higher threshold cutoff that limited IgG detection sensitivity compared to the spike protein. This reactivity to RBD may be driven by artificial epitopes, exposure of epitopes otherwise inaccessible within the context of the native-like S glycoprotein trimer or a product of microsphere coupling.

Immunoassay detection of IgG antibodies that can bind to RBD has been used as a surrogate for neutralization tests which require cell-culture, pseudoviruses, or biosafety-containment and wild-type SARS-CoV-2 (25, 28, 29). SARS-CoV-2 neutralizing antibodies target the S glycoprotein S1 subunit, particularly the RBD and N-terminal domains, and sterically interfere with human ACE-2 receptor interaction (30-34). Yet, SARS-CoV-2 and MERS-CoV neutralizing monoclonal antibodies have been identified that binds epitopes that do not interfere with receptor engagement (35, 36). Furthermore, non-neutralizing antibody-mediated protection has been observed in other virus infections, including HIV and Ebola virus (37, 38). Given the relatively poor performance of this RBD protein in this β-CoV MMIA, and our ability to capture the full-breadth of the humoral response, e.g., RBD-binding, neutralizing and non-neutralizing, to SARS-CoV-2 infection with the native-like spike protein trimer (39), future studies will exclude the monomeric RBD antigen from β-CoV MMIA strategy.

The MMIA approach provided additional antibody detection data that is suggestive of SARS-CoV-2 IgG cross-reactivity with SARS-CoV-1 and MERS-CoV. Conserved cross-neutralizing epitopes between MERS-CoV, SARS-CoV-1 and SARS-CoV-2 S glycoproteins have been identified (35, 40). Whether SARS-CoV-2 induced polyclonal IgG antibody responses to SARS-CoV-1 and MERS-CoV spike proteins are retained after affinity maturation, or are cross-neutralizing requires further investigation. Future studies with this MMIA will incorporate spike proteins from the seasonal alphacoronaviruses, HCoV-NL63 and HCoV-229E, to improve upon specificity for SARS-CoV-2 antibody detection. We hypothesize that the inclusion of NL63 and 229E spike proteins will provide additional off-target control of cross-reactive antibodies to SARS-CoV-2 spike protein, decreasing the threshold cutoff for positive SARS-CoV-2 and enabling improved detection of waning and low positive SARS-CoV-2 IgG antibody. To attempt to achieve similar success at detecting SARS-CoV-2 antibodies and seroprevalence four months after diagnosis (41), next steps in MMIA development will be focused on a re-calibration of threshold cutoffs using archived sera in an MMIA that is limited to spike proteins from SARS-CoV-2, HCoV-HKU1, HCoV-OC43, HCoV-NL63 and HCoV-229E. Importantly, when utilized in select subject cohorts from prospective, longitudinal observational studies in which serum samples are obtained before SARS-CoV-2 infection, this MMIA approach has the potential to measure antibody cross reactions with seasonal HCoVs, and investigate whether HCoV-induced antibodies confer any protection against COVID-19.

## CONCLUSION

In summary, we have presented the development of a multiplex microsphere-based immunoassay for SARS-CoV-2 serology that includes envelope spike glycoproteins from zoonotic, SARS-CoV-1 and MERS-CoV, and seasonal endemic betacoronaviruses, HCoV-HKU1 and HCoV-OC43. Performance assessment of this immunoassay with serum samples from a pre-2019 archived sera bank and SARS-CoV-2 PCR positive subjects who sought medical treatment at military hospitals demonstrated 100% specificity for SARS-CoV-2 IgG antibody detection and 98% sensitivity with samples collected as early as 10 days after the onset of symptoms. Through this multiplex approach we are able to measure the potential cross reactions of HCoV-induced antibodies to SARS-CoV-2 spike glycoproteins. Application of this multiplex approach to prospective observational studies will enable the direct examination of whether pre-existing HCoV-induced antibodies affect COVID-19 clinical outcomes, i.e. asymptomatic presentation and symptomatic severity.

## MATERIALS AND METHODS

### Recombinant protein antigens and microsphere coupling

Prefusion stabilized SARS-CoV-2 S-2P glycoprotein ectodomain trimers (hereafter referred to as spike protein) and SARS-CoV-2 RBD were purchased from LakePharma, Inc. (Hopkinton, MA USA). This SARS-CoV-2 spike protein shares an equivalent ectodomain with the NIH Vaccine Research Center designed SARS-CoV-2 S-2P protein, and the Mount Sinai SARS-CoV-2 S-2P protein used in ELISA-based serology (26, 27, 42-44). Differences between LakePharna, Inc. and VRC or Mount Sinai spike protein constructs are highlighted in the C-terminus tags and selection of mammalian cell-line for expression.

Design and expression of prefusion stabilized HCoV-HKU1, HCoV-OC43, SARS-CoV-1 and MERS-CoV spike proteins have been previously described (15, 42). A mock antigen, consisting of cell culture supernatant from untransfected HEK cells was collected via centrifugation then filtered through a 0.22 µM PES filter to remove debris. Mock antigen-coupled beads are included in each microtiter well to control for non-specific/artificial antisera binding; samples that react with the mock antigen above an established 3-fold cutoff are retested. Spike proteins were coupled to carboxylated magnetic MagPlex microspheres (Bio-Rad, Hercules, CA) at a protein to microsphere ratio of 15 µg:100 µL, and antigen-coupled microspheres were resuspended in a final volume of 650 µL following manufacturer’s protocol (Bio-Rad) for amine coupling.

### Non-human primate sera

Archived sera were used from rhesus macaques inoculated with a total dose of 2.6×10^6^ TCID50 of SARS-CoV-2 via a combination of intranasal, intratracheal, oral and ocular inoculation routes (16). Serum samples were collected at dpi 0 (baseline), 1, 3, 5, 7, 10, 12, 14, 17 and 21. To purify serum IgG antibody, 250 µL of serum from each of four experimentally infected NHPs collected 21 dpi were pooled then subjected to thermal inactivated for 30 minutes at 60 °C. During inactivation, 2 mL of Protein G agarose, 50% suspension (Sigma-Aldrich, St. Louis, Missouri, USA) was added to a chromatography column and the buffer was allowed to flow through. The bead bed was then washed three times with 10 mL of PBS. Inactivated pooled sera were diluted 1:5 in PBS then added to the column. Flow-through was collected, then re-added to the column; this process was repeated for a total of three passes through the column. The bead bed was again washed three times with 10 mL of PBS. Finally, IgG was eluted from the Protein G agarose using a 0.1M Glycine elution buffer, pH 2.5 then returned to neutral pH using 1M Tris-HCl, pH 8.0. Eluted IgG was concentrated using an Amicon Ultra Centrifugal Filter unit (Merck Millipore, Burlington, Massachusetts, USA), and the buffer was exchanged to a 1X PBS buffer containing 25% glycerol.

SARS-CoV-2 IgG and IgM antibody seroconversion was determined as the first dpi where a 4-fold increase in the median fluorescence intensity (MFI) was measured compared to the baseline sera collection. Between the 1:250 and 1:1000 dilutions, some NHP IgG antibody reactivities were no longer saturating the upper level of the MMIA. We chose further sera screening at a 1:400 dilution, retaining the ability of the MMIA to detect positives at near MMIA saturation, i.e. >20,000 MFI, while still within the linear region of detection.

### Participant enrollment and sera collection

SARS-CoV-2 negative human serum specimens utilized were from archived sera collected between 2012 – 2018 in the Infectious Disease Clinical Research Program (IDCRP) Acute Respiratory Infection Consortium Natural History Study (ARIC, IDCRP-045) (45). ARIC sera predate the COVID-19 pandemic and were collected from subjects who had nasopharyngeal swabs tested by nucleic acid amplification methods for virus etiologies of acute respiratory infections; samples collected from individuals with rhinovirus and the seasonal human coronaviruses HCoV-OC43, -HKU1, -229E and -NL63 were used (46). In addition, serum samples were collected since the emergence of SARS-CoV-2 under the IDCRP Epidemiology, Immunology, and Clinical Characteristics of Emerging Infectious Diseases with Pandemic Potential (EPICC, IDCRP-085) protocol; a prospective, longitudinal study to analyze COVID-19 disease. Subjects were enrolled at five hospitals across the continental U.S., including Walter Reed National Military Medical Center (WRNMMC, Bethesda, MD), Brooke Army Medical Center (BAMC, San Antonio, TX), Naval Medical Center San Diego (NMCSD, San Diego, CA), Madigan Army Medical Center (MAMC, Tacoma, WA) and Fort Belvoir Community Hospital (FBCH, Fort Belvoir, VA). Subjects of all race and gender seeking treatment for acute illness at these military hospitals were offered enrollment into the EPICC, IDCRP-085 protocol. Study enrollment included subjects with laboratory-confirmed SARS-CoV-2 infection by nucleic acid amplification test, subjects with compatible illness in whom SARS-CoV-2 infection is initially suspected but PCR confirmed as SARS-CoV-2 negative, and asymptomatic subjects at risk of SARS-CoV-2 due to high risk exposure. In this study, 422 sera samples from 204 individual subjects were tested. The earliest serum samples collected ≥ 10 dpso from subjects with longitudinal samples were included in positive performance agreement. Additionally, serum samples from 35 subjects undergoing treatment at the COVID-19 field hospital at the Jacob K. Javits Convention Center (New York, NY) under the COVID-19 Antibody Prevalence in Military Personnel Deployed to New York (COVID-19 NYC) protocol were included in the assessment of assay performance. EPICC, IDCRP-085 and COVID-19 NYC protocols were approved by the Uniformed Services University Institutional Review Board.

### Multiplex microsphere-based immunoassay screening procedures

Serum samples were collected from venipuncture in serum separator tubes, processed and stored at -80 °C in 250 µL aliquots until use. For each 96-well plate, a multiplex master mix of antigen-coupled microspheres was made by diluting 100 µL of each antigen-coupled microsphere working stock into 10 mL (1:100) 1XPBS without calcium and magnesium (Corning Inc., Corning, NY) (all mentions of PBS refer to solutions without calcium and magnesium), and 100 µL of this master mix were added to each well so that each well contained 1 µL (∼23 ng) of each antigen-coupled microsphere per well. Wells were washed with 1XPBS + 0.05% Tween20 + 0.02% sodium azide two times. One hundred microliters of each serum sample was added to each well. Serum samples were initially diluted within a class II type A2 biological safety cabinet (BSC) then subjected to thermal inactivation for 30 min at 60 °C, further serum dilutions are noted in each respective figure legend. Human serum samples (1.25 µL) were diluted 1:400 in PBS and tested in technical duplicate A and B plates. Controls on each duplicate plate included a PBS blank (wells: A1, B1, G12, H12) and positive (C1, F12) and negative (D1, E12) non-human primate serum. As testing progressed, PCR and serology confirmed human positive/negative samples replaced non-human primate serum samples as the qualified controls for inter- and intra-plate variation.

Samples were incubated at room temperature for 45 minutes with agitation (900 rpm), and plates were washed three times. Secondary antibody (goat anti-human IgG cross-absorbed biotin-conjugated or goat anti-human IgM cross-absorbed biotin-conjugated; Thermo Fisher Scientific, Waltham, MA) was diluted 1:5000 in 1XPBS + 0.05% Tween20 (PBST) and 100 µL of each secondary was added to each well and incubated for 45 minutes with agitation, and plates were washed three times. Streptavidin-phycoerythrin (Bio-Rad) was diluted 1:1000 in PBST and 100 µL was then added to each well and incubated for 30 minutes with agitation, and plates were washed three times. Lastly, 100 µL of PBST was added to each well and plates were resuspended by agitation for 5 minutes. Plates were read on Bio-Plex 200 multiplexing systems (Bio-Rad) with PMT voltage setting to the High RP1 target and 100 bead count requirements. The MFI for the four PBS blank wells on each plate were subtracted from the MFI of each sample well and MFI values for samples are reported as the PBS adjusted average from duplicate plates.

### Threshold cutoffs for SARS-CoV-2 antibody

To establish threshold cutoffs for SARS-CoV-2 spike protein-specific antibody reactivity, we tested 127 archival acute and convalescent human serum samples from ARIC. Acute and convalescent serum samples were collected within approximately three and twenty-eight days of symptom onset, respectively. A cut-off of three times the mean MFI obtained using a mock antigen preparation coupled microsphere was initially used to determine positivity of cross-reactive antibodies in archival serum samples. As cross-reactive antibodies were found to occur in 4 out of 45 serum samples from archival HCoV PCR-positive individuals, we then established a cut-off of three standard deviations above the mean (99.7% probability) MFI of these archival HCoV convalescent serum samples (n= 43) to establish a positivity threshold for detection of SARS-CoV-2 spike protein reactive IgG and IgM antibodies. The remaining 84 archival serum samples were tested against this MFI threshold cutoff for SARS-CoV-2 reactivity. The 127 archival ARIC serum samples were tested in technical duplicates in three independent experiments to establish threshold cutoffs and specificity for SARS-CoV-2.

### Enzyme-linked immunosorbent assay

Flat bottom 96-well microtiter plates (Corning) were coated with 300 ng of SARS-CoV-2 spike protein per well diluted in 100 µl of ELISA coating buffer (1XPBS, 5.3g Na_2_CO_3_, 4.2g NaHCO_3_, pH 9.6) and incubated overnight at 4 °C. The next day, spike protein was removed and 125 µl of 5% BSA blocking buffer were added to each well and incubated for 1 hour at 37 °C. SARS-CoV-2 spike protein coated and blocked plates were then wasted plate three times with 200 µl PBST. Serum samples were subjected to thermal inactivation after being initially diluted in a BSC. Inactivated serum samples were then serially diluted 2-fold in PBS. One hundred microliters of each dilution was added in duplicate to the antigen coated plate, sealed, and incubated at 37°C for 1 hour. Plates were then washed three times with 200 µl PBST. One hundred microliters of secondary antibody, anti-human (H&L) HRP conjugated, diluted 1:5000 in PBS to each well was added to each well and plates were incubated at 37°C for 1 hour. Plates were then washed three times with 200 µl PBST. Eighty-five microliters ABST Substrate Solution (Thermo Fisher Scientific) was added to each well and plates were agitated (900 rpm) at room temperature for 30 minutes, then analyzed at 650 nm absorbance on a plate reader (Molecular Devices, San Jose, CA).

### Statistical analysis

Figures were generated and statistical analyses were performed in GraphPad Prism version 7.0. The positive predictive value and negative predictive value were calculated with MedCalc statistical software. ROC analysis was conducted using R version 4.0.2.

## Data Availability

The protocols and datasets used and/or analyzed during the current study are available from the corresponding author(s) on reasonable request.

## DECLARATIONS

These research protocols, IDCRP-085, IDCRP-045 and COVID-19 NYC, were approved by the USU IRB.

## DISCLAIMER

The contents of this publication are the sole responsibility of the author(s) and do not necessarily reflect the views, opinions, or policies of the Uniformed Services University (USU), the Henry M. Jackson Foundation for the Advancement of Military Medicine, Inc. (HJF), National Institutes of Health or the Department of Health and Human Services, Brooke Army Medical Center, the U.S. Army Medical Department, the U.S. Army Office of the Surgeon General, the US Department of Defense (DoD), the Departments of the Air Force, Army or Navy, or the U.S. Government. Mention of trade names, commercial products, or organization does not imply endorsement by the U.S. Government. A number of the co-authors are military service members (or employees of the U.S. Government). This work was prepared as part of their official duties. Title 17 U.S.C. §105 provides that ‘Copyright protection under this title is not available for any work of the United States Government.’ Title 17 U.S.C. §101 defines a U.S. Government work as a work prepared by a military service member or employee of the U.S. Government as part of that person’s official duties.

## ACKNOWLEDGEMENTS

We thank Kelly Snead, Vanessa Wall, John-Paul Denson, Simon Messing, and William Gillette (Protein Expression Lab, FNCLR) for excellent technical assistance. We also thank Scott Merritt and Katrin Mende (IDCRP, HJF, Brooke Army Medical Center) and Kathleen Pratt (Department of Medicine, USUHS) for assistance with sample acquisition.

## REFERENCES

1. E. Petersen et al., Comparing SARS-CoV-2 with SARS-CoV and influenza pandemics. Lancet Infect Dis 20, e238–e244 (2020).

2. A. Wu et al., Genome Composition and Divergence of the Novel Coronavirus (2019- nCoV) Originating in China. Cell Host Microbe 27, 325–328 (2020).

3. F. J. Ibarrondo et al., Rapid Decay of Anti-SARS-CoV-2 Antibodies in Persons with Mild Covid-19. N Engl J Med 10.1056/NEJMc2025179 (2020).

4. Q. X. Long et al., Clinical and immunological assessment of asymptomatic SARS-CoV-2 infections. Nat Med 26, 1200–1204 (2020).

5. J. Seow et al., Longitudinal evaluation and decline of antibody responses in SARS-CoV-2 infection. medRxiv 10.1101/2020.07.09.20148429, 2020.2007.2009.20148429 (2020).

6. E. Hartenian et al., The molecular virology of Coronaviruses. J Biol Chem 10.1074/jbc.REV120.013930 (2020).

7. U.S. Food and Drug Administration (2020, September 11) EUA authorized serology test performance. https://www.fda.gov/medical-devices/coronavirus-disease-2019-covid-19-emergency-use-authorizations-medical-devices/eua-authorized-serology-test-performance

8. A. Ayouba et al., Development of a Sensitive and Specific Serological Assay Based on Luminex Technology for Detection of Antibodies to Zaire Ebola Virus. J Clin Microbiol 55, 165–176 (2017).

9. R. L. Powell et al., A Multiplex Microsphere-Based Immunoassay Increases the Sensitivity of SIV-Specific Antibody Detection in Serum Samples and Mucosal Specimens Collected from Rhesus Macaques Infected with SIVmac239. Biores Open Access 2, 171–178 (2013).

10. N. G. Satterly, M. A. Voorhees, A. D. Ames, R. J. Schoepp, Comparison of MagPix Assays and Enzyme-Linked Immunosorbent Assay for Detection of Hemorrhagic Fever Viruses. J Clin Microbiol 55, 68–78 (2017).

11. R. Kozak et al., Severity of coronavirus respiratory tract infections in adults admitted to acute care in Toronto, Ontario. J Clin Virol 126, 104338 (2020).

12. S. Nickbakhsh et al., Extensive multiplex PCR diagnostics reveal new insights into the epidemiology of viral respiratory infections. Epidemiol Infect 144, 2064–2076 (2016).

13. S. Su et al., Epidemiology, Genetic Recombination, and Pathogenesis of Coronaviruses. Trends Microbiol 24, 490–502 (2016).

14. B. Freeman et al., Validation of a SARS-CoV-2 spike protein ELISA for use in contact investigations and serosurveillance. bioRxiv : the preprint server for biology 10.1101/2020.04.24.057323, 2020.2004.2024.057323 (2020).

15. J. Hicks et al., Serologic cross-reactivity of SARS-CoV-2 with endemic and seasonal Betacoronaviruses. medRxiv 10.1101/2020.06.22.20137695 (2020).

16. V. J. Munster et al., Respiratory disease in rhesus macaques inoculated with SARS-CoV-2. Nature 10.1038/s41586-020-2324-7 (2020).

17. W. Deng et al., Primary exposure to SARS-CoV-2 protects against reinfection in rhesus macaques. Science 369, 818–823 (2020).

18. S. Lu et al., Comparison of nonhuman primates identified the suitable model for COVID-Signal Transduct Target Ther 5, 157 (2020).

19. C. Shan et al., Infection with novel coronavirus (SARS-CoV-2) causes pneumonia in Rhesus macaques. Cell Res 30, 670–677 (2020).

20. M. Norman et al., Ultrasensitive high-resolution profiling of early seroconversion in patients with COVID-19. Nature Biomedical Engineering 10.1038/s41551-020-00611-x (2020).

21. L. Liu et al., A preliminary study on serological assay for severe acute respiratory syndrome coronavirus 2 (SARS-CoV-2) in 238 admitted hospital patients. Microbes and infection 22, 206–211 (2020).

22. B. Lou et al., Serology characteristics of SARS-CoV-2 infection after exposure and post-symptom onset. European Respiratory Journal 56, 2000763 (2020).

23. L. Guo et al., Profiling Early Humoral Response to Diagnose Novel Coronavirus Disease (COVID-19). Clinical Infectious Diseases 71, 778–785 (2020).

24. N. M. A. Okba et al., Severe Acute Respiratory Syndrome Coronavirus 2-Specific Antibody Responses in Coronavirus Disease Patients. Emerging Infectious Disease journal 26, 1478 (2020).

25. L. Premkumar et al., The receptor-binding domain of the viral spike protein is an immunodominant and highly specific target of antibodies in SARS-CoV-2 patients. Science Immunology 5, eabc8413 (2020).

26. F. Amanat et al., A serological assay to detect SARS-CoV-2 seroconversion in humans. Nat Med 26, 1033–1036 (2020).

27. C. Klumpp-Thomas et al., Standardization of enzyme-linked immunosorbent assays for serosurveys of the SARS-CoV-2 pandemic using clinical and at-home blood sampling. medRxiv 10.1101/2020.05.21.20109280 (2020).

28. F. Wu et al., Neutralizing antibody responses to SARS-CoV-2 in a COVID-19 recovered patient cohort and their implications. medRxiv 10.1101/2020.03.30.20047365, 2020.2003.2030.20047365 (2020).

29. C. W. Tan et al., A SARS-CoV-2 surrogate virus neutralization test based on antibody-mediated blockage of ACE2-spike protein-protein interaction. Nat Biotechnol 38, 1073–1078 (2020).

30. L. Liu et al., Potent neutralizing antibodies against multiple epitopes on SARS-CoV-2 spike. Nature 584, 450–456 (2020).

31. C. O. Barnes et al., Structures of Human Antibodies Bound to SARS-CoV-2 Spike Reveal Common Epitopes and Recurrent Features of Antibodies. Cell 182, 828-842.e816 (2020).

32. X. Chi et al., A neutralizing human antibody binds to the N-terminal domain of the Spike protein of SARS-CoV-2. Science 369, 650–655 (2020).

33. D. F. Robbiani et al., Convergent antibody responses to SARS-CoV-2 in convalescent individuals. Nature 584, 437–442 (2020).

34. P. J. M. Brouwer et al., Potent neutralizing antibodies from COVID-19 patients define multiple targets of vulnerability. Science 369, 643–650 (2020).

35. D. Pinto et al., Cross-neutralization of SARS-CoV-2 by a human monoclonal SARS-CoV antibody. Nature 583, 290–295 (2020).

36. L. Wang et al., Importance of Neutralizing Monoclonal Antibodies Targeting Multiple Antigenic Sites on the Middle East Respiratory Syndrome Coronavirus Spike Glycoprotein To Avoid Neutralization Escape. Journal of Virology 92, e02002–02017 (2018).

37. L. M. Mayr, B. Su, C. Moog, Non-Neutralizing Antibodies Directed against HIV and Their Functions. Frontiers in immunology 8, 1590–1590 (2017).

38. E. O. Saphire, S. L. Schendel, B. M. Gunn, J. C. Milligan, G. Alter, Antibody-mediated protection against Ebola virus. Nat Immunol 19, 1169–1178 (2018).

39. S. J. Zost et al., Potently neutralizing and protective human antibodies against SARS-CoV-2. Nature 584, 443–449 (2020).

40. J. Zang et al., Immunization with the receptor-binding domain of SARS-CoV-2 elicits antibodies cross-neutralizing SARS-CoV-2 and SARS-CoV without antibody-dependent enhancement. Cell Discov 6, 61–61 (2020).

41. D. F. Gudbjartsson et al., Spread of SARS-CoV-2 in the Icelandic Population. N Engl J Med 382, 2302–2315 (2020).

42. D. Esposito et al., Optimizing high-yield production of SARS-CoV-2 soluble spike trimers for serology assays. bioRxiv 10.1101/2020.05.27.120204 (2020).

43. R. N. Kirchdoerfer et al., Stabilized coronavirus spikes are resistant to conformational changes induced by receptor recognition or proteolysis. Sci Rep 8, 15701 (2018).

44. D. Wrapp et al., Cryo-EM structure of the 2019-nCoV spike in the prefusion conformation. Science 367, 1260–1263 (2020).

45. C. Coles, E. V. Millar, T. Burgess, M. G. Ottolini, The Acute Respiratory Infection Consortium: A Multi-Site, Multi-Disciplinary Clinical Research Network in the Department of Defense. Mil Med 184, 44–50 (2019).

46. M. Bouvier et al., Species-specific clinical characteristics of human coronavirus infection among otherwise healthy adolescents and adults. Influenza Other Respir Viruses 12, 299–303 (2018).

